# Syncope and COVID-19 disease – a systematic review

**DOI:** 10.1101/2020.12.30.20249060

**Authors:** Raquel Falcão de Freitas, Sofia Cardoso Torres, José Pedro L. Nunes

## Abstract

**Background:** Syncope is not a common manifestation of COVID-19, but it may occur in this context and it can be the presenting symptom in some cases. Although several mechanisms may explain the pathophysiology behind COVID-19 related syncope, a valid relationship has not been established yet. In this systematic review, we aimed to examine the current incidence of syncope in COVID-19 patients and to explore different patterns observed in this setting.

**Methods:** A systematic review across PubMed, ISI Web of Knowledge and SCOPUS was performed, according to PRISMA guidelines, in order to identify all relevant articles regarding both COVID-19 and syncope.

**Results:** We identified 81 publications, of which 62 were excluded. The cumulative incidence of syncope and pre-syncope across the selected studies was 7.1% (256/3584 patients). Unspecified syncope was the most common type (76.2% of the reported episodes), followed by reflex syncope (18.1% of the cases). Orthostatic hypotension was responsible for 3.6% of the cases and syncope of presumable cardiac cause accounted for 2.0%. Arterial hypertension was present in 64.7% of the patients and either angiotensin receptor blockers or angiotensin converting enzyme inhibitors were used by 39.5% of hypertensive patients with syncope.

**Conclusion:** Syncope, although not considered a typical symptom of the COVID-19 disease, can be associated with it, particularly in early stages. Different types of syncope were seen in this context, each with different implications requiring distinct approaches. A careful reevaluation of blood pressure whenever a patient develops COVID-19 is suggested, including reassessment of antihypertensive therapy.

## INTRODUCTION

The ongoing Coronavirus pandemic has proved to be a challenging setback to the health of the world population, since its first cases were announced in the city of Wuhan, China, around December, 2019. Millions of cases and a considerable number of deaths have been reported worldwide.

It is currently known that, although the novel SARS-CoV-2 virus often leads to significant disease in the respiratory system, it can also negatively affect several other vital organ systems. Significant damage, namely, to the cardiovascular, nervous and hematopoietic systems has been outlined and an impact in hemostasis has also been thoroughly discussed as blood hypercoagulability is common among hospitalized COVID-19 patients. (1) Regarding the cardiovascular manifestations, heart failure, thromboembolism, myocarditis, arrhythmias, pericarditis and acute coronary syndromes have been described in this context. (2, 3). On the other hand, the most common neurological symptoms reported in COVID-19 have been smell and taste disturbances, headache, myalgia, and altered mental status. (4)

Syncope is largely defined as a transient loss of conscience (TLOC) associated with cerebral hypo-perfusion. In the light of a severe systemic disease, non-traumatic TLOC can have distinct etiologies, varying from the benign reflex syncope and syncope due to orthostatic hypotension to the increasingly serious cardiac syncope. (5) Apart from unspecified syncope, these three main groups stem from different mechanisms and, therefore, may require specialized treatment. Consequently, an accurate diagnosis becomes imperative.

Recently, some case-series have emerged reporting syncope as a possible symptom of the COVID-19 infection, whether it developed at the onset or during the course of the disease. (3) It is important to mention that some of these reports outline its occurrence days before the main respiratory symptoms. (6) If a valid relationship between COVID-19 and syncope is established, a number of patients could be isolated in a timely manner, minimizing the contagious phase. In the present report, we aimed to systematically review the current published literature that correlates syncope with COVID-19.

## METHODS

### Search Strategy

A comprehensive literature search was carried out with the purpose of identifying all reported articles relating syncope to COVID-19, according to the guidelines for Preferred Reporting Items for *Systematic Reviews and Meta-Analyses*. This search was conducted on the databases Medline (PUBMED), ISI Web of Knowledge and SCOPUS.

The search query, which took place on the 14-26 October 2020, included the following MeSH terms and keywords: “COVID-19 AND syncope”; “SARS-COV-2 AND syncope”. Additionally, we scanned the list of references from the included studies in this analysis.

### Inclusion and Exclusion Criteria

Studies were included if they simultaneously described COVID-19 disease and syncope, presented as a possible symptom of the infection. We included case-series, case-reports, cross-sectional studies with prospective data collection and retrospective analyses.

Articles that were not available in English (2 reports) and publications with no original data were excluded. We also excluded articles that did not describe the clinical characteristics of the patients or of the episode of syncope in a significant way.

### Study Eligibility Assessment

Study eligibility was individually assessed by two investigators. Considering the significant amount of case reports found, no formal quality assessment was conducted. We found it unfeasible to conduct an appropriate meta-analysis, mainly due to limited research data among studies as well as visible discrepancies across results.

### Data Extraction

The investigators individually assessed whether the studies addressed the topic in question and if all the inclusion/exclusion criteria were met. Initially, this was done according to the “screening phase”, where only the title and the abstract were analyzed. This was followed by the “inclusion phase”, where the integral text was evaluated.

## RESULTS

### Study Selection

With the use of the first set of keywords, we obtained 26 results from Medline, 8 from ISI Web of Knowledge and 21 from SCOPUS (Figure 1). From the second set of keywords, 12 articles from Medline, 5 from ISI Web of Knowledge and 9 from SCOPUS were extracted – with a total number of 19 articles selected for the purpose of the present study (Figure 1). The complete set of selected studies is presented in Table 1. SARS-COV-2 infection was diagnosed by real-time reverse transcriptase polymerase chain reaction (RT-PCR) or a chest X-ray or CT scan showing the characteristic bilateral interstitial pneumonia of COVID-19 in all cases, except in the report by Romero-Sánchez et al., in which a minority of patients were diagnosed by means of serological testing. (7)

**Table 1.** Summary of included articles. Pts - patients; yo - year old; ARBs - angiotensin receptor blockers; ICU - intensive care unit; PPM - permanent pacemaker implantation; ECG - electrocardiogram; ICD - implantable cardioverter-defibrillator; AV - atrioventricular; ACE-I - angiotensin-converting-enzyme inhibitors; CMR - cardiac magnetic resonance; EEG - electroencephalogram; CSF - cerebrospinal fluid; CT-computed tomography; MRI - magnetic resonance imaging; BMI - body mass index; RT-PCR - real time polymerase chain reaction. For references see text.

| Source | Study Type | Country | Patients (n) | Patient Characteristics | Comorbidities | Main Findings | Syncope Type |
| --- | --- | --- | --- | --- | --- | --- | --- |
| Oates *et al.* | Retrospective Study | United States of America | 77<br>(study group: 37 pts (32 with syncope and 5 with presyncope); control group: 40 pts without syncope/presyncope) | Study group:<br>Median age: 69<br>51% males<br>24% Caucasian | Study group:<br>- Hypertension (68%)<br>- Obesity (42%)<br>- Diabetes Mellitus (32%)<br>- Coronary Artery Disease (27%)<br>- Atrial Fibrillation (8%) | - Incidence of syncope/presyncope was 3.7%<br>- Study group: greater use of ARBs (p=0.03)<br>- Systolic blood pressure lower in the study group (p=0.01)<br>- Pulse rate lower in study group (p<0.0001)<br>- Study group required less ICU admissions & had lower need for mechanical ventilation | - 59.4% Unspecified<br>- 15.6% Neurocardiogenic/Reflex<br>- 12.5% Hypotensive<br>- 3.1% Cardiopulmonary |
| Chen *et al.* | Prospective Study | United States of America | 102<br>(study group: 24 pts with syncope, near syncope or nonmechanical fall) | Study group:<br>Mean age: 61 | Study group:<br>- Cardiovascular disease history in 29% | - More pts from the study group required oxygen, had gastrointestinal symptoms and elevated troponin levels compared to the rest of the pts (p>0.05). | - Unspecified |
| Ebrille *et al.* | Case-series | Italy | 5<br>(2 pts with syncope and 3 with presyncope) | Patient 1: 71 yo male<br>Patient 2: 65 yo female<br>Patient 3: 79 yo male<br>Patient 4: 75 yo male<br>Patient 5: 75 yo male | Patient 1: Hypertension, Coronary Artery Disease and PPM<br>Patient 2: Mitral Valve stenosis, PPM for AV block, Atrial Fibrillation<br>Patient 3: Hypertension, Diabetes Mellitus, Transient Ischemic Attack, 3<sup>rd</sup> degree AV block<br>Patient 4: Chagas disease, PPM due to AV block, ICD because of ventricular tachycardia<br>Patient 5: Dilated Cardiomyopathy, PPM | - All pts had an episode of syncope as the only initial symptom of COVID-19 infection<br>- 4/5 pts were on ACE-I chronic therapy<br>Syncope due to arrhythmia, structural cardiac disease or pulmonary embolism was excluded. | - Reflex Syncope<br>- Autonomic Dysfunction, either primary or secondary |
| Chang *et al.* | Case Report | United States of America | 1 | 49 yo | No significant medical | - Syncope and fever | - Presumable Cardiac Cause |
|  |  |  |  | Male | history | - ECG with Brugada pattern, uncovered during fever<br>- Brugada Syndrome diagnosis |  |
| Pasquetto *et al.* | Case Report | Italy | 1 | 52 yo<br>Male | - | - Dyspnea and fever<br>- 2 episodes of syncope during high fever<br>- The ECG presented a “coved-type” aspect in leads V1 and V2 and a first-degree AV block<br>- Brugada Syndrome diagnosis | - Presumable Cardiac Cause |
| Luetkens *et al.* | Case Report | Germany | 1 | 79 yo<br>Male | Asthma | - Elevated troponin and NT-proBNP<br>- CMR showed diffuse interstitial myocardial edema with mild systolic dysfunction and mild pericardial effusion.<br>- COVID-19 associated myocarditis diagnosis | - Presumable Cardiac Cause |
| Logmin *et al.* | Case Report | Germany | 1 | 70 yo<br>Female<br>History of syncope over the years but not lately | - Psoriatic Arthritis<br>- Neuropathic Pain<br>- Paroxysmal Atrial Fibrillation | - 3 syncopes, one of which convulsive.<br>- Respiratory symptoms emerged later<br>- Normal long-term ECG, blood pressure monitoring and Schellong test<br>- Brain MRI without acute alterations (signs of minimal previous ischaemic events)<br>- Normal EEG and CSF analysis<br>- Pathological Sympathetic Skin Response | - Autonomic Dysfunction |
| Tapé *et al.* | Case Report | United States of America | 1 | 79 yo<br>Female | - Coronary Artery Disease with multiple stents<br>- Hypertension<br>- Congestive Heart Failure | - Syncopal episode, myalgias, cough and fever<br>- Initially admitted in the Emergency Department for a syncopal workup<br>- BP of 116/62 mm Hg sitting & 85/50 mm Hg standing<br>- Normal ECG and telemetry monitoring<br>- Fever later prompted SARS-CoV-2 testing<br>- Developed respiratory failure later | - Orthostatic Hypotension or Reflex Syncope |
| Canetta *et al.* | Retrospective Analysis | Italy | 103<br>(35 pts with syncope and 68 pts without) | Mean age<br>With syncope: 74<br>Without syncope: 72 | Syncope group:<br>- Hypertension (45.7%)<br>- Dyslipidemia (17%)<br>- Renal insufficiency (20%)<br>- Hypothyroidism (5.7%) | - Hypocapnic Hypoxemia in most pts<br>- Mean heart rate was significantly lower in pts who experienced syncope<br>- Pts reporting syncope had a normal cardiac assessment | - Reflex Syncope |
|  |  |  |  | 69% of the pts with syncope were male | - Dementia (11.4%)<br>- Cancer (11.4%)<br>- Atrial Fibrillation (5.7%) |  |  |
| Singhania *et al.* | Case Report | United States of America | 1 | 71 yo<br>Female | Hypertension | -Syncope and later altered mental status as the only complaints that ultimately led to COVID-19 diagnosis.<br>-Tachycardic, normal ECG | - Orthostatic Hypotension |
| Sang *et al.* | Case Report | United States of America | 1 | 62 yo<br>Male | - Hypertension<br>- Dyslipidemia | -Patient with syncope, ventricular fibrillation and shock secondary to a massive pulmonary embolism in the setting of SARS-CoV-2 infection | - Cardiopulmonary Syncope |
| Birlutiu *et al.* | Case-series | Romania | 4 | Patient 1: 67 yo Caucasian male<br>Patient 2: 65 yo Caucasian female<br>Patient 3: 61 yo Caucasian male<br>Patient 4: 48 yo Caucasian female | Patient 1: Hypertension, Stroke, Diabetes Mellitus<br>Patient 2: Diabetes Mellitus, hypertension, 3<sup>rd</sup> degree AV block with pacemaker<br>Patient 3: -<br>Patient 4: - | - All 4 pts presented with syncope after micturition.<br>- 2 of them had associated intense, persistent headaches either preceding or post syncope and 1 of them had diffuse abdominal pain and nausea as warning signs.<br>- Time of syncope varied from the 2<sup>nd</sup> day to the 11<sup>th</sup> day of hospitalization and 2 pts had repeated syncope over a 2-minute interval and suffered acute traumatic brain injury as a consequence.<br>-Cardiologic investigation was normal in all 4 pts and there was no evidence of hypotension. | Reflex Syncope |
| Huang *et al.* | Case Report | United States of America | 1 | 40 yo<br>Female | - Diabetes Mellitus<br>- Obesity | - Patient presented with fever & syncope and was admitted for encephalitis<br>- CSF was later found to be positive for SARS-COV-2 on reverse transcription polymerase chain reaction | Probable Autonomic Dysfunction |
| Radmanesh *et al.* | Retrospective Study | United States of America | 242<br>(79 of whom presented with syncope) | Mean age of 68.7<br>62% male<br>38% female | - | - 242 pts underwent at least 1 brain imaging (CT and/or MRI) examination within 2 weeks of testing positive for COVID-19<br>- Syncope/fall was one of the most common clinical indications for imaging (79 pts, 32.6%)<br>- Of the 13 pts with acute/subacute infarcts, 2 were imaged due to syncope/fall.<br>- Among the 7 pts with acute hemorrhage, the clinical indication was syncope in 4 pts. | - Unspecified |
| Chachkhiani *et al.* | Retrospective | United States | 250 | Mean age of 60 | Group with Neurological | - 34 (14%) pts had a neurological chief complaint at | - Unspecified |
|  | Study | of America | (6 of whom had syncope) | 45% male<br>80% African American | chief complaint:<br>Hypertension (71%)<br>Diabetes (44%) | presentation, and syncope was one of the most common complaints (2%)<br>- Neurological complaints at presentation and during the hospital stay are associated with a higher risk of death, prolonged hospital stay, and intubation |  |
| García-Monco *et al.* | Cross-sectional study with prospective data | Spain | 35<br>(2 of whom had syncope) | Median Age: 66<br>71% male | Hypertension (57%)<br>Dyslipidemia (60%)<br>Diabetes Mellitus (17%)<br>Smoking (23%)<br>Obesity (BMI ≥30) (6%) | - Pts who presented with or developed a neurological disorder and were diagnosed with COVID-19 were analyzed. Of the 35 pts, 2 had syncope.<br>- The CSF of one of these pts with syncope was tested for the presence of SARS-Cov-2 using the RT-PCR assay and was negative. | - Unspecified |
| Xiong *et al.* | Retrospective Cohort study | China | 917<br>(3 of whom had syncope) | Mean age: 48.7<br>55% male | 44% had non-neurologic comorbidities<br>3% had neurologic comorbidities | - Of the 917 people with COVID-19, 39 had new-onset neurologic events (3 with syncope).<br>- The pts with syncope were three women aged between 52 and 61 years without a previous history of neurologic or systemic disorders. ECGs recorded afterwards were normal. - Brain CT conducted in one of the syncope pts did not reveal any new lesions. | - Unspecified |
| Romero-Sánchez *et al.* | Retrospective, observational study | Spain | 841<br>(5 of whom had syncope) | Mean age: 66.42<br>56% male | Hypertension<br>Dyslipidemia<br>Obesity<br>Heart disease<br>Diabetes Mellitus<br>Chronic kidney disease | - Of 841 pts hospitalized with COVID-19, 57.4% developed some form of neurologic symptom.<br>- 5 pts suffered from a syncopal episode, all of which had non-severe COVID-19 disease.<br>- In 21 pts (2.5%), a neurologic manifestation was the reason that prompted the visit to the emergency department. 2 of those pts presented with syncope. | - Unspecified |
| Argenziano *et al.* | Retrospective Case-Series | United States of America | 1000<br>(48 of whom had syncope) | Median age: 63.0<br>66.9% male | Hypertension (60%)<br>Diabetes Mellitus (37%) | - Syncope was the presenting symptom in:<br>4% of pts in whom the highest level of care was the emergency department;<br>5.7% of pts who needed hospital care;<br>3% of pts who needed care in the ICU | - Unspecified |

**Figure 1.**
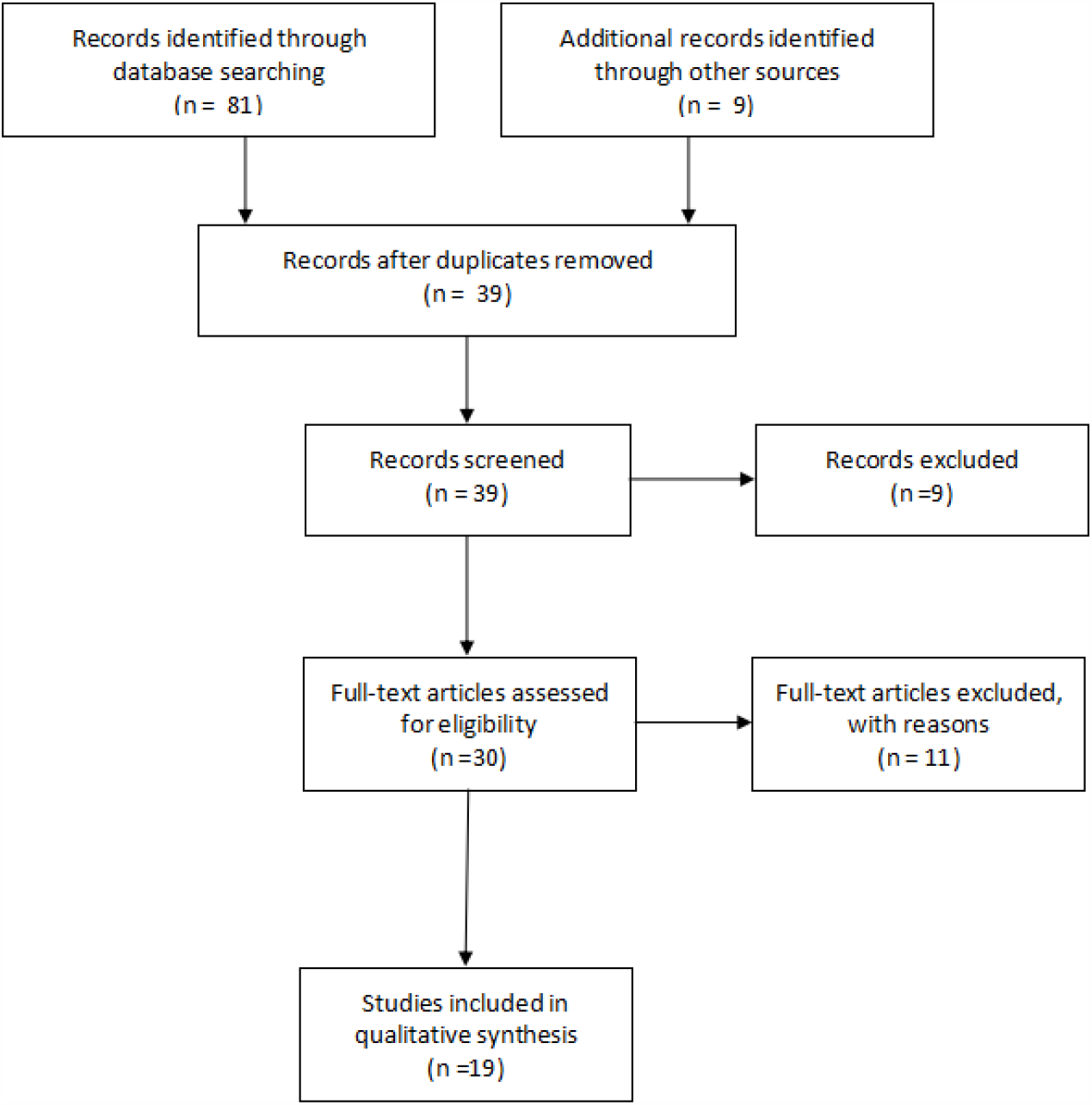
Flowchart showing literature search method. n = number of articles.

The excluded reports are presented in Table 2, with the corresponding reasons.

**Table 2.**
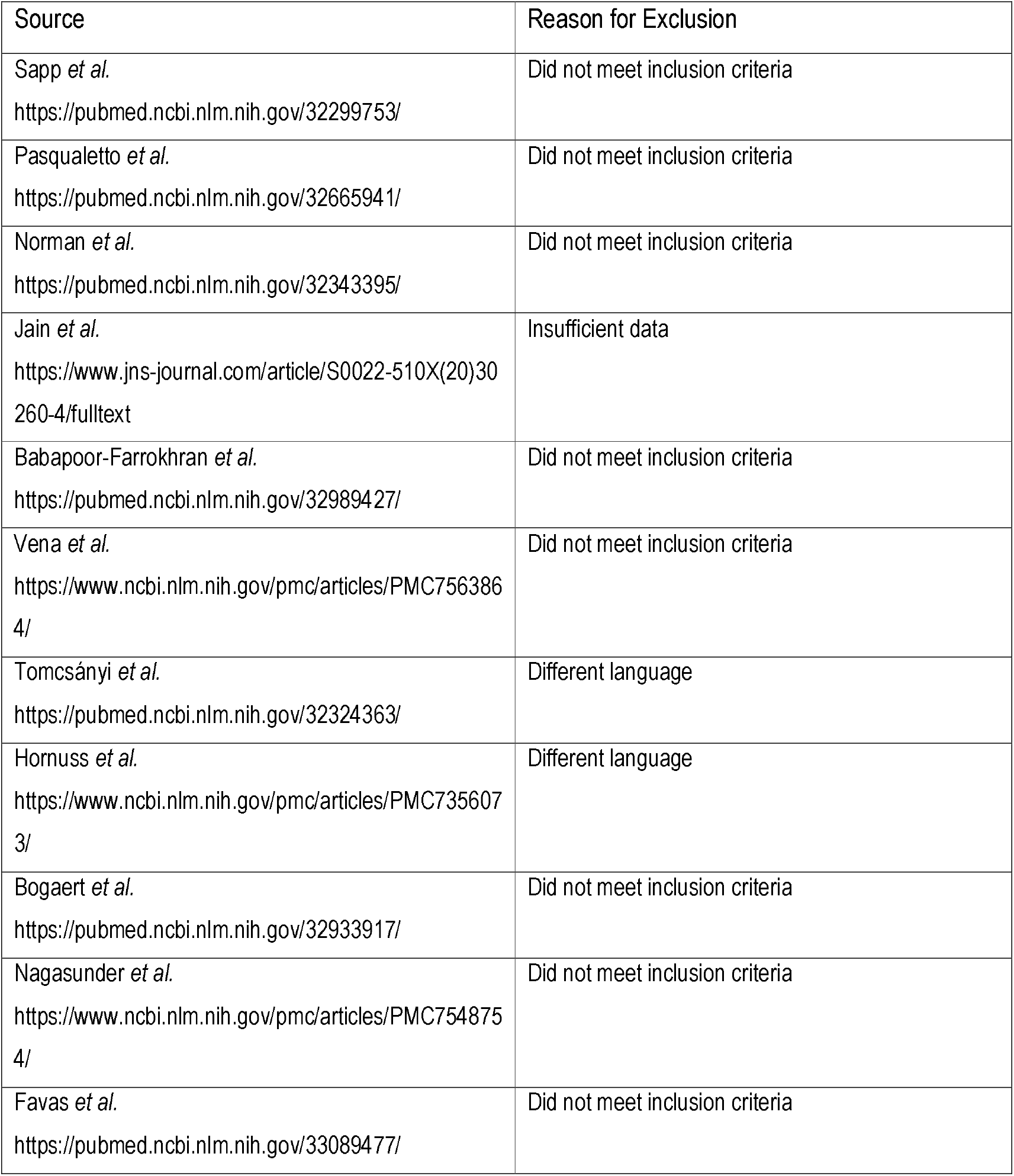
Articles excluded, with reasons

| Source | Reason for Exclusion |
| --- | --- |
| Sapp *et al.*<br>https://pubmed.ncbi.nlm.nih.gov/32299753/ | Did not meet inclusion criteria |
| Pasqualetto *et al.*<br>https://pubmed.ncbi.nlm.nih.gov/32665941/ | Did not meet inclusion criteria |
| Norman *et al.*<br>https://pubmed.ncbi.nlm.nih.gov/32343395/ | Did not meet inclusion criteria |
| Jain *et al.*<br>[https://www.jns-journal.com/article/S0022-510X(20)30260-4/fulltext](https://www.jns-journal.com/article/S0022-510X%2820%2930260-4/fulltext) | Insufficient data |
| Babapoor-Farrokhran *et al.*<br>https://pubmed.ncbi.nlm.nih.gov/32989427/ | Did not meet inclusion criteria |
| Vena *et al.*<br>https://www.ncbi.nlm.nih.gov/pmc/articles/PMC7563864/ | Did not meet inclusion criteria |
| Tomcsányi *et al.*<br>https://pubmed.ncbi.nlm.nih.gov/32324363/ | Different language |
| Hornuss *et al.*<br>https://www.ncbi.nlm.nih.gov/pmc/articles/PMC7356073/ | Different language |
| Bogaert *et al.*<br>https://pubmed.ncbi.nlm.nih.gov/32933917/ | Did not meet inclusion criteria |
| Nagasunder *et al.*<br>https://www.ncbi.nlm.nih.gov/pmc/articles/PMC7548754/ | Did not meet inclusion criteria |
| Favas *et al.*<br>https://pubmed.ncbi.nlm.nih.gov/33089477/ | Did not meet inclusion criteria |

### Study Characteristics

Of the 19 included articles, 8 were case reports (8) (9) (10) (11) (12) (13) (14) (15), 6 were retrospective analyses (3) (7) (16) (17) (18) (19), 3 were case-series (6) (20) (21) and 2 were prospective studies (22) (23). Given the nature of the topic at hand, every article was published in 2020. The United States of America contributed with most of the studies, with a total of 10 articles. Eight articles were conducted in Europe and one was from China.

The size of the included studies varied from 1 to 1000 patients. The mean age ranged from 40 to 79 years of age. Considering the studies in which it was possible to extrapolate gender information of the patients who experienced syncope, we found that 60% were men.

### Synthesis of Results

There were 256 cases of syncope and pre-syncope (248 with syncope and 8 with pre-syncope) contemplated in this review, comprising a relatively low percentage of the total number of COVID-19 infections.

The majority of studies, 8, described syncopal episodes of unspecified etiology. (3) (7) (16) (17) (18) (21) (22) (23) Six studies attributed the cause to orthostatic hypotension (3) (6) (8) (12) (14) (15), while another 5 studies alluded to possible cardiac syncope (3) (9) (10) (11) (13). In turn, reflex syncope was highlighted in 4 of the studies. (3) (6) (19) (20)

As stated before, unspecified syncope was the most common cause of the transient loss of conscience, translating into 76.2% (189/248) of the reported episodes.

The overall relative incidence of reflex syncope was 18.1% (45/248). Orthostatic hypotension accounted for 3.6% (9/248) and presumable cardiac syncope was responsible for 2.0% (5/248). Regarding comorbidities, hypertension was heavily represented in 11 studies (3) (6) (7) (8) (9) (14) (16) (19) (20) (21) (22), making it the most prevalent comorbid condition exhibited by the participants. This was closely followed by Diabetes mellitus, obesity, dyslipidemia and heart disease. As shown in Table 3, detailed data concerning comorbidities were available for 88 patients, and data concerning drug usage in patients with arterial hypertension were available for 76 patients. Arterial hypertension was present in 64.7% of patients and either angiotensin receptor blockers or angiotensin converting enzyme inhibitors were used by 39.5% of hypertensive patients with COVID-19 and syncope (Table 3).

**Table 3.** Associated clinical conditions in 88 patients with COVID-19 and syncope and drug usage in 76 hypertensive patients with COVID-19 and syncope

| **<u>Associated conditions</u>** | **<u>Medications</u>** | **<u>N (%)</u>** |
| --- | --- | --- |
| **Arterial hypertension** |  | 64.7%<br>(57/88) |
|  | Angiotensin receptor blockers OR<br>angiotensin converting enzyme inhibitors | 39.5%<br>(30/76) |
|  | Diuretics | - |
|  | Beta blockers | 34.2%<br>(26/76) |
|  | Calcium channel blockers | 14.5%<br>(11/76) |
| **Diabetes mellitus** |  | 18.2%<br>(16/88) |
| **Obesity** |  | 18.2%<br>(16/88) |
| **Coronary Heart Disease** |  | 12.5%<br>(11/88) |
| **Dyslipidemia** |  | 8.0% (7/88) |
| **Atrial Fibrillation** |  | 8.0% (7/88) |
| **Renal Insufficiency** |  | 8.0% (7/88) |

Out of the 8 case reports, syncope was the reason that prompted the visit to the Emergency Department in 7 of them (87.5%). (8) (10) (11) (12) (13) (14) (15) Furthermore, Ebrille *et al.* (6) and *Canetta et al.* (19) also described syncope as the presenting symptom of the infection. On the other hand, Argenziano *et al*. (21) stated that syncope was the presenting symptom in 4% of patients in whom the highest level of care was the emergency department, 5.7% of patients who required hospital care and 3% of patients who needed care in the ICU.

Considering confirmed or presumable cardiac syncope, the 5 reported cases included situations of myocarditis (11), massive pulmonary embolism (PE) and ventricular fibrillation (9), Brugada Syndrome (13) (10) and new onset atrial fibrillation together with anterior wall ST elevation MI (3) as the underlying conditions.

The most common laboratory findings included elevated C-reactive protein and elevated troponin. Lymphocytopenia was also characteristic.

## DISCUSSION

The results of our review suggest that syncope is a relatively uncommon manifestation of the COVID-19 infection, however, when it does occur, it is often the presenting symptom and should warrant further investigation. The timely association of syncope with COVID-19 disease could, in some cases, offer an opportunity to mitigate the dissemination of the disease by means of an early diagnosis. Risk stratification and treatment could perhaps also improve as a consequence.

### Reflex Syncope

There are two main pathophysiological mechanisms involved in reflex syncope, both of them relying on an imbalance between sympathetic and parasympathetic activity leading to an inappropriate reflex. These are vasodepression caused by insufficient sympathetic vasoconstriction and cardioinhibition due to parasympathetic predominance. (6)

The mechanism that precipitates this type of syncope in the context of COVID-19 remains unknown, but it has been speculated that it could be coupled with the affinity of SARS-Cov-2 to the ACE-2 receptors, promoting their internalisation and affecting the baroreflex and chemoreceptor responses. (19) Two studies (6) (19) highlighted the fact that the study group consistently showed a lower mean heart rate in comparison to the control group. These observations could support the hypothesis stated before, that the inappropriate baroreflex response would lead to a partial inhibition of the compensatory increase of heart rate during acute hypoxemia. Since a significant amount of patients who experienced syncope also suffered some degree of hypoxia, it is possible that these two conditions are mechanistically related. (3)

Moreover, in the report by Oates et al. (3), the study cohort displayed a lower heart rate and lower systolic blood pressures at admission, as well as lower intensive care unit requirement, which, in turn, suggests that the syncope observed may not have been associated with increased incidence of severe COVID-19 infection.

Arterial hypertension was present in almost two thirds of patients with COVID-19 and syncope for whom detailed data were available. This may indicate that maintaining the usual antihypertensive medication may be inadequate in some cases of COVID-19. The present results suggest a careful reevaluation of blood pressure whenever a hypertensive patient develops COVID-19.

Some studies take notice of a greater use of angiotensin receptor blocking agents (3) (6) among the patients who suffered from syncopal episodes. An explanation for this might be the upregulation of ACE-2 expression in many tissues leading to a facilitated binding of the virus to the cells. On the other hand, the blood pressure lowering effects caused by the inhibition of the vasopressor effect of angiotensin II, carried out by these agents, could also play an additional role in the occurrence of syncope.

### Orthostatic Hypotension

Autonomic dysfunction frequently plays an important role in the etiology of orthostatic hypotension. In the context of COVID-19, this might arise from a primary failure caused by the virus itself or a secondary failure due to autoimmune autonomic neuropathy. (6) It is known that vascular injury caused by IL-1 and IL-6, in addition to decreased systemic vascular resistance resulting in vasodilation, could be one of the possible explanations for the occurrence of syncope. (6)

Syncope may indeed precede the appearance of COVID-19 typical symptoms. Interestingly, one patient (8) who was initially admitted in the Emergency Department for a syncopal workup and in whom the syncopal episode was attributed to orthostatic hypotension, had, at the time of presentation, a normal chest radiograph and no findings suggestive of infection other than lymphopenia. She was later diagnosed with COVID-19 and developed respiratory failure. This emphasizes the importance of identifying syncope as a possible atypical sign of infection so that an early diagnosis can be established.

The presence of neurologic features in COVID-19 infection has also been described, for example, under the form of stroke (24), polyneuropathy (25) and Guillain-Barré syndrome. (26) A study included in this review outlined an infection completely confined to the central nervous system, with no involvement of other organ systems. (15) In that case, the patient presented with fever and syncope and was later diagnosed with COVID-19. The respective cerebrospinal fluid (CSF) analysis was found to be positive for SARS-CoV-2 on reverse transcriptase polymerase chain reaction and the case was considered consistent with SARS-CoV-2 encephalitis. However, determining if the encephalitis was caused by a direct effect of the virus or if it was due to a critical autoimmune reaction or inflammation owing to a cytokine storm remains questionable.

### Cardiac Syncope

This type of syncope can develop from either arrhythmias or structural damage to the heart and great vessels. The five conditions that were linked to cardiac syncope in this review have been stated before, and they are known to be frequently associated to abnormal ECG findings. Therefore, baseline ECG should be granted to every COVID-19 patient presenting with, or, having a history of, syncope.

The ECG of the pulmonary embolism patient showed anterior ST segment elevations and subsequently (after ventricular fibrillation and resuscitation) displayed a wide complex rhythm consistent with ventricular tachycardia, with right bundle branch block and left axis deviation. (9) The overall prevalence of venous thromboembolism in the setting of COVID-19 is poorly defined, with current case series suggesting an approximate value between 20.6–25%. (27) Regarding Brugada syndrome, two patients (10, 13) presented with syncope, fever and Brugada pattern on the 12-lead ECG, which led to the diagnosis of this syndrome and resulted in the implantation of a subcutaneous implantable cardioverter defibrillator (S-ICD) in one patient and use of a wearable cardioverter defibrillator in the other. Fever is known to unmask Brugada’s ECG pattern and to precipitate ventricular arrhythmias in these patients and one study showed that more than half of the study cohort experienced syncope or cardiac arrest in the setting of a fever (28). Since fever is one of the most characteristic COVID-19 symptoms, it is reasonable to expect that previously unrecognized Brugada syndromes may appear during this pandemic.

Lastly, in the report by Luetkens et al. (11), the patient presented with fatigue, shortness of breath and recurrent syncopes. His ECG was normal. The elevated high-sensitivity troponin and the presence of diffuse intersticial myocardial edema with mild systolic dysfunction on cardiac magnetic resonance (CMR) imaging led to the diagnosis of COVID-19 associated myocarditis. The diffuse myocardial inflammation could be related to direct damage of cardiomyocytes by the virus or myocardial injury triggered by a cytokine storm and imbalanced response by type 1 and type 2 T-helper cells. This patient had an atypical presentation of COVID-19 disease, with no fever and only mild symptoms of pneumonia, but, instead, with progressive cardiac involvement and diffuse myocardial inflammation. Recovery was observed on follow-up, after treatment was initiated for heart failure.

### Limitations

The majority of studies in the present review are case reports, hence, the limitations include those intrinsic to this kind of studies. There is a lack of ability to generalize the results, mainly due to the fact that there are no large epidemiological data available. In addition, since causality cannot be definitively inferred from an uncontrolled observation, we cannot be completely certain that the occurrence of syncope was related to the COVID-19 infection. We also analyzed a substantial amount of retrospective studies, most of which were not designed to primarily assess the incidence of syncope in the context of COVID-19 infection. We attempted at making a distinction between the different types of syncope, however, in some studies, we found it impossible to do so. Regarding the cardiac syncope cases, we assumed that the TLOC had most likely a cardiac origin, nevertheless, the data available were not enough to be sure in some cases. Furthermore, our exclusion criteria limited the articles to those written in the English language and that may have omitted two eligible studies.

## Conclusions

Syncope is an uncommon symptom of the COVID-19 infection and there is limited evidence regarding this association. Out of all the types of syncope, cardiac syncope was the less incident in this review (2%). Nevertheless, the presence of syncope should always call for special attention in order to prevent serious complications.

It is important to correlate syncope with COVID-19, since a significant amount of studies showed that it can be the presenting symptom. Being able to recognize this unusual presentation can lead to an earlier diagnosis, meaning that we could be one step closer to controlling the dissemination of this pandemic.

Arterial hypertension was particularly prevalent in patients with COVID-19 and syncope, which may indicate that maintaining the usual antihypertensive medication may be inadequate in some cases of COVID-19. A careful reevaluation of blood pressure whenever a patient develops COVID-19 is suggested, including reassessment of antihypertensive therapy. Larger, more directed, multicenter epidemiological studies are required in order to truly evaluate the incidence, etiology and impact of syncope in this group of patients.

## Data Availability

Data obtained from the published scientific literature

## Notes

### Competing Interest Statement

The authors have declared no competing interest.

### Funding Statement

No external funding received

### Author Declarations

No ethics committee approval is needed for systematic reviews

